# Medical Diagnosis Coding Automation: Similarity Search vs. Generative AI

**DOI:** 10.1101/2024.04.26.24306470

**Authors:** Vanessa Klotzman

**Author notes:** Contributing authors.

## Abstract

**Objective:** This study aims to predict ICD-10-CM codes for medical diagnoses from short diagnosis descriptions and compare two distinct approaches: similarity search and using a generative model with few-shot learning.

**Materials and Methods:** The text-embedding-ada-002 model was used to embed textual descriptions of 2023 ICD-10-CM diagnosis codes, provided by the Centers provided for Medicare & Medicaid Services. GPT-4 used few-shot learning. Both models underwent performance testing on 666 data points from the eICU Collaborative Research Database.

**Results:** The text-embedding-ada-002 model successfully identified the relevant code from a set of similar codes 80% of the time, while GPT-4 achieved a 50 % accuracy in predicting the correct code.

**Discussion:** The work implies that text-embedding-ada-002 could automate medical coding better than GPT-4, highlighting potential limitations of generative language models for complicated tasks like this.

**Conclusion:** The research shows that text-embedding-ada-002 outperforms GPT-4 in medical coding, highlighting embedding models’ usefulness in the domain of medical coding.

## 1 Background and Significance

The International Classification of Disease (ICD), established by the World Health Organization (WHO) is the universally recognized and standardized system for medical coding worldwide. It provides a comprehensive framework for categorizing diseases, health conditions, and related information, facilitating accurate and consistent documentation, data sharing, and research across the global healthcare community. It is employed by healthcare providers worldwide to categorize diseases and conditions. Medical coding involves the assignment of ICD codes like ICD-10-CM codes to classify diagnoses and reasons for visits in all healthcare settings, is essential for guiding clinical decisions, tracking diseases, and impacting healthcare financing[1, 2].

Medical coding is traditionally manual, with coders translating physicians’ notes into the appropriate ICD codes while adhering to complex guidelines. In this process, highly trained medical coders assign ICD (International Classification of Diseases) codes to patient encounters based on the information found in clinicians’ notes, however, manual ICD coding is time-consuming and error-prone, making the quality and productivity of coding a matter of concern in practice. The process is error-prone [3–9] due to the complexity of medical language and coding guidelines. Coders often need help with subtle differences between disease subtypes, leading to misclassification. Physicians’ use of abbreviations and synonyms which adds to the ambiguity [10, 11]. Making this a non-trivial task for humans. Furthermore, inexperienced coders may incorrectly assign separate codes to related diagnoses, a problem called unbundling, which can result in costly mistakes [12]. These coding inaccuracies have substantial financial implications, contributing to an estimated annual expenditure of $25 billion in the United States, as reported by Lang et al. [13] Farkas et al. [14]. With recent AI technologies (e.g., NLP), automated medical coding has the potential to support clinical coders better.

Automated Medical Coding (AMC) is the idea that artificial intelligence can automate clinical coding. In recent years, there has been a significant increase in AMC-related work [14–32] through deep learning. Although research in this field has grown, this problem is far from being solved [18, 33].For instance, automated coding remains a complex problem because extracting knowledge from patients’ clinical records is challenging. These records are not uniformly structured, the medical field’s terminologies can be complicated for non-professionals to comprehend, and physicians often have different ways of describing symptoms, leading to various descriptions for the same disease.

Embeddings [34] in Natural Language Processing (NLP) represent words as real-valued vectors. These vectors can capture the meaning of words in such a way that words closer together in the vector space are expected to have similar meanings. In clinical NLP [35], embeddings are helpful for analyzing medical data and texts, aiding decision-making and research. The use of word embeddings in Automated Medical Coding (AMC) systems [36–43] is increasingly being explored as it has the potential to bridge the gap between the informal language of medical diagnoses and the formal language of ICD code descriptions. For instance CAIC [27] uses cross-textual attention to match parts of medical notes with ICD codes. While GatedCNN-NCI [18] creates a network linking every aspect of medical notes to ICD codes. BiCapsNetLE [44] integrates ICD code descriptions into word embeddings of clinical notes, enhancing alignment. DLAC [45] employs a description-based label attention mechanism, focusing on the correlation between the descriptions of ICD codes and the features of medical notes. ICDBigBird uses a Graph Convolutional Network (GCN) and ehances the ICD code emebddings by using their relational structure.

Even though, there is a growing body of work for utilizing embeddings in clinical coding, there has been a growing interest of what a Large Language Model(LLM) can do in the health sector due to their ability of understanding, generating, and predicting new content.

As the interest in Large Language Models (LLMs) continues to grow in the health sector, as evidenced by multiple recent studies studies [46–57], our objective is to compare the effectiveness of two distinct approaches to predict ICD-10-CM codes accurately. We will compare the effectiveness of similarity search, for which we will be using text-embedding-ada-002 [58], and an LLM, in which we will be using GPT-4 [59] from OpenAI to predict ICD-10-CM codes.

## 2 Materials and Methods

### 2.1 Data collection

We utilized the diagnosis strings (patient diagnoses) from the eICU Collaborative Research Database [60], which contains data from different critical care units (CCUs) across the United States from patients who were admitted between 2014 and 2015. We selected a subset of 666 patients from the total dataset of 2,710,672 patients. This sample size represents a 99% confidence level with a 5% margin of error [61]. We utilize each patient’s current diagnoses from the data we collected, which comprise of the diagnosis string and the corresponding ICD-10 CM codes. The diagnosis strings will serve as inputs to the models, with the ICD-10-CM codes as the outputs. The ICD-10-CM codes will be used for comparison to assess the model’s accuracy in prediction. The diagnosis strings in the eICU database are organized in a tiered system. For example, “neurologic—trauma - CNS—intracranial injury—with subarachnoid hemorrhage” shows this: it starts with a general category “neurologic”, goes into a more specific “trauma - CNS”, then to “intracranial injury”, and ends with a detailed aspect “with subarachnoid hemorrhage”. Each part of the string represents a deeper level of diagnosis detail. Table 1, shows sample diagnosis strings and their corresponding ICD-10 CM codes from the dataset we are using.

**Table 1.** eICU Sample Data: Diagnosis & ICD-10 Code.

| Diagnosis | ICD-10 CM Code |
| --- | --- |
| burns/trauma\|dermatology\|cellulitis | L03.90 |
| burns/trauma\|trauma - chest\|lung trauma | S27.30 |
| hematology\|coagulation disorders\|DVT | I80.9 |

### 2.2 Model Selection

We utilize OpenAI’s text-embedding-ada-002 model ^1^, as it surpasses previous models in text search and text similarity from OpenAI. We evaluate the effectiveness of the embedding model relative to the latest version of GPT-4, which we operated using the Microsoft OpenAI Azure Service. Our selection of only the text-embedding-ada-002 model and GPT-4 was due to the constraints set by the terms and conditions of using the PhysioNet dataset ^2^.

### 2.3 text-embedding-ada-002

In this work, we utilized the text-embedding-ada-002 model to embed the textual descriptions of 2023 ICD-10-CM diagnosis codes ^3^, as provided by the Centers for Medicare & Medicaid Services source. After generating these embeddings, our primary objective was to evaluate their performance. To do so, we used the dataset of diagnosis strings obtained from the eICU dataset.

To assess the accuracy of matching medical diagnoses (diagnosis strings) with their respective ICD-10-CM codes, we inputted these diagnosis strings into the text-embedding-ada-002 model. Our objective was to determine if this single model could accurately return the closest ICD-10-CM code based on the ICD-10 description. Additionally, the model returned the top four ICD-10-CM codes for each medical condition. We selected four as the default value for similarity searches by vector, because this is specified as the standard setting in the LangChain documentation ^4^. The workflow of how the embeddings function is clearly illustrated in Figure 1 The embeddings are generated and then stored in the vector database. These embeddings correspond to the textual descriptions of the 2023 ICD-10-CM diagnosis codes. When a user submits a query as a medical diagnosis (referred to as the diagnosis string), the system searches the database for embeddings similar to the embedding of the query. Finally, the system retrieves the closest ICD-10-CM codes based on the similarity between embeddings, providing relevant matches for the medical diagnosis.

**Fig. 1.**
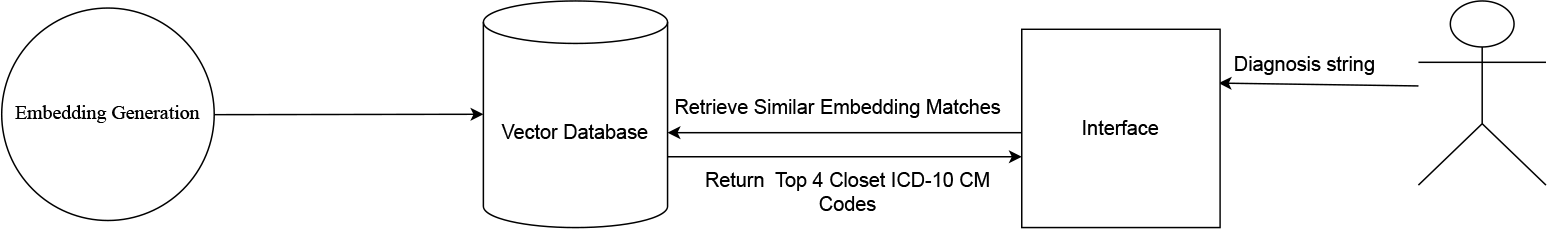
Visualizing Automated ICD-10 Code Prediction Process: Streamlining Medical Coding

### 2.4 GPT-4

We prompted GPT-4 with few-shot prompting to assess its capability in medical coding. Few shot prompting [62] was selected because large language models have notable zero-shot abilities, but they tend to perform poorly in complex tasks when using zero-shot settings. Few-shot prompting serves as a method to enable in-context learning, where demonstrations in the prompt help direct the model toward better performance. Figure 2 contains the prompt we used. The examples for the prompt were acquired by clustering the diagnosis strings. We used K-means clustering [63] to group our diagnosis strings and found that 8 clusters worked best. The ideal number of eight clusters was determined using the elbow method [64], which evaluates the within-cluster sum of squares across a range of 1 to 20 possible clusters; the ‘elbow’ point, where there is a significant decrease in within-cluster dissimilarity, indicates the most suitable number of clusters. We picked a range between 1 and 20 as it is a manageable number of clusters that can be effectively interpreted and analyzed. From each cluster, we selected the diagnosis closest to the cluster’s center in the vector space as the most representative example of the cluster; these representative examples were then used in the few-shot prompt.

**Fig. 2.**
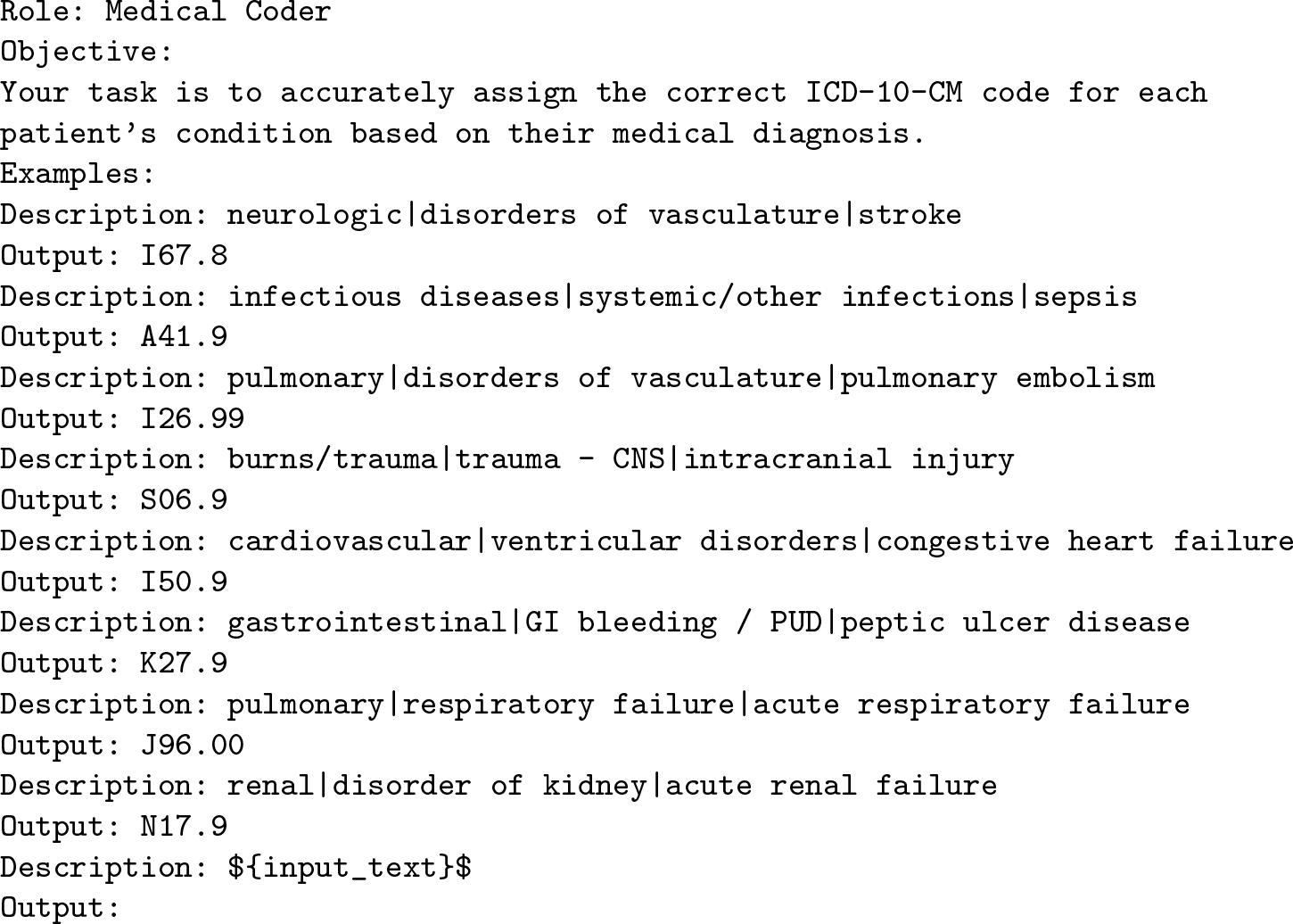
GPT-4 Prompt

To optimize GPT-4 for medical coding, we experimented with different temperature settings (0.1, 0.5, 0.9) on the sample of 666 diagnoses collected for this study, representing a 99% confidence interval and a 5% margin of error. Our results showed that a temperature of 0.1 was effective, as it balanced the model’s creative outputs and the need for accurate, deterministic responses in medical coding.

## 3 Results

The text-embedding-ada-002 model achieved an 80% accuracy rate in identifying the correct ICD-10-CM codes from the retrieved similar codes, outperforming GPT-4, which achieved a 50% accuracy rate in the same task. This suggests that embedding models, like text-embedding-ada-002, can offer improved accuracy and efficiency in medical coding.

## 4 Discussion

In this study, we discovered that embedding models like text-embedding-ada-002 could potentially be more effective than GPT-4, a large language model. The critical advantage of embeddings lies in their focus on the semantic similarity of words, an aspect vital for accurately matching medical diagnoses with ICD codes, as this technique allows for a more precise understanding and interpretation of medical terminology, which is crucial in medical coding. Embeddings analyze the context and meaning of words more concentratedly, leading to higher accuracy in identifying relevant codes.

Moreover, when assessing the feasibility of using embedding models like text-embedding-ada-002, it becomes evident that these models align well with medical coding requirements. They offer a more focused approach, potentially assisting in accurately linking diagnoses with the correct ICD codes, which demands precision. It suggests that embedding models better fit medical coding tasks compared to more generative models like GPT-4, which handle a broader range of data.

In contrast, GPT-4 processes a wide range of data and contexts. While this versatility is helpful for general tasks, it can lead to less precision in specialized areas like medical coding, where specific terminology and accurate coding are essential. GPT-4’s handling of vast information might make it more challenging to differentiate between similar medical terms and codes, potentially affecting its performance in this field.

## 5 Conclusion

The results indicate that embedding models like text-embedding-ada-002 appear more suitable for medical coding tasks than large language models like GPT-4. This result could be primarily due to text-embedding-ada-002’s focused approach on the semantic similarity of words, which has led to an 80% accuracy in identifying ICD-10-CM codes, significantly surpassing GPT-4’s 50% accuracy. Embedding models like text-embedding-ada-002 are a more practical choice for medical coding due to their precision in analyzing and understanding medical terminology. On the other hand, GPT-4, although capable of broad data processing, may prove less effective in specialized fields such as medical coding, where accuracy and specific terminology are crucial. Hence, for precision-dependent tasks such as medical coding, embedding models like text-embedding-ada-002 could offer a more suitable solution than generative models like GPT-4.

## Data Availability

All data produced in the present study are available upon reasonable request to the authors.

## Footnotes

1 https://openai.com/blog/new-and-improved-embedding-model

2 https://physionet.org/news/post/415

3 https://www.cms.gov/medicare/coding-billing/icd-10-codes

4 https://api.python.langchain.com/

